# COVID-19 risk score as a public health tool to guide targeted testing: A demonstration study in Qatar

**DOI:** 10.1101/2021.03.06.21252601

**Authors:** Laith J. Abu-Raddad, Soha Dargham, Hiam Chemaitelly, Peter Coyle, Zaina Al Kanaani, Einas Al Kuwari, Adeel A. Butt, Andrew Jeremijenko, Anvar Hassan Kaleeckal, Ali Nizar Latif, Riyazuddin Mohammad Shaik, Hanan F. Abdul Rahim, Gheyath K. Nasrallah, Hadi M. Yassine, Mohamed G. Al Kuwari, Hamad Eid Al Romaihi, Mohamed H. Al-Thani, Abdullatif Al Khal, Roberto Bertollini

## Abstract

**Background:** The objective of this study was to develop a Coronavirus Disease 2019 (COVID-19) risk score to guide targeted RT-PCR testing in Qatar.

**Methods:** The Qatar national COVID-19 testing database was analyzed. This database includes a total of 2,688,232 RT-PCR tests conducted between February 5, 2020-January 27, 2021. Logistic regression analyses were implemented to identify predictors of infection and to derive the COVID-19 risk score, as a tool to identify those at highest risk of having the infection. Score cut-off was determined using the receiving operating characteristic (ROC) curve based on maximum sum of sensitivity and specificity. The score’s performance diagnostics were assessed.

**Results:** Logistic regression analysis identified age, sex, and nationality as significant predictors of infection and were included in the risk score. The score’s scoring points were lower for females compared to males and higher for specific nationalities. The ROC curve was generated and the area under the curve was estimated at 0.63 (95% CI: 0.63-0.63). The score had a sensitivity of 59.4% (95% CI: 59.1%-59.7%), specificity of 61.1% (95% CI: 61.1%-61.2%), a positive predictive value of 10.9% (95% CI: 10.8%-10.9%), and a negative predictive value of 94.9% (94.9%-95.0%). The risk score derived early in the epidemic, based on data until only April 21, 2020, had a performance comparable to that of a score based on a year-long testing.

**Conclusions:** The concept and utility of a COVID-19 risk score were demonstrated in Qatar. Such a public health tool, based on a set of non-invasive and easily captured variables can have considerable utility in optimizing testing and suppressing infection transmission, while maximizing efficiency and use of available resources.

## Introduction

Suppressing the severe acute respiratory syndrome coronavirus 2 (SARS-CoV-2) epidemic necessitates strategic preparedness and response [1]. The World Health Organization (WHO) has urged countries to adopt a “testing, tracing, and isolation” approach as the “backbone” of their SARS-CoV-2 national response [2]. However, to suppress the epidemic, deliver healthcare services to those in need, and ensure optimal use of resources, testing strategies need to be guided by real-time data analysis so that testing is prioritized to those at higher risk of exposure.

A risk score is an objective set of simple questions or measurements that can be used to assess the likelihood of an individual having a specific infection/disease condition [3-6]. Such scores have been useful in designing initial screening or testing strategies for a variety of diseases, as they reduce the need for more invasive, time-consuming, and expensive testing, while optimizing resource allocation by targeting individuals at higher risk of having the infection/disease [7]. The utility of developing a risk score for SARS-CoV-2 infection offers the benefits of earlier case detection, isolation of cases, and quarantine of contacts, given the disease burden associated with this infection.

Qatar is a high-income country in the Arabian Gulf with a total population of 2.8 million, the majority of whom (88%) are expatriates from over 150 countries [8-10]. The nation’s rapid development resulted in a unique socio-demographic structure dominated by men, who comprise 74% of the total population [8], and by younger age cohorts (ages 20-50 years), who likewise comprise 74% of the population [8].

The country was afflicted with a large first epidemic wave of SARS-CoV-2 infection that peaked toward the end of May, 2020 [11]. As of January 29, 2020, >65,000 infections per million population had been laboratory-confirmed [11-13]. Qatar has also one of the world’s most extensive databases to document this epidemic and its toll at the national level [14], such that Qatar’s epidemic has been one of the most thoroughly investigated and best characterized [11, 14-24].

This study had three objectives. The first was to present a derived risk score for SARS-CoV-2 infection that was developed during the first epidemic wave in April, 2020 to inform the national response to the epidemic. The second objective was to assess the prospective performance of this risk sore on epidemic data collected after its derivation. The third objective was to update this risk score to end of January, 2021, and to assess its diagnostic metrics for future use as part of the national response.

The overarching goal of this study was to demonstrate the feasibility and utility of the concept of a Coronavirus Disease 2019 (COVID-19) risk score as a public health tool in an emergent epidemic, applying it to a specific country. Building on the public health utility of risk scores for other diseases such as diabetes [3-6], we believe that this study provides the first COVID-19 risk score for any country. The score has been named “*COVID-19 risk score”*, given the prevailing public use of “COVID-19”, as opposed to SARS-CoV-2.

## Methods

### Data source

We analyzed the national database for SARS-CoV-2 real-time polymerase chain reaction (RT-PCR) testing compiled by Hamad Medical Corporation (HMC), the main public healthcare provider in Qatar. The database includes results of all RT-PCR testing conducted in Qatar, regardless of whether it was for suspected SARS-CoV-2 cases, traced contacts, infection surveillance, or other purposes, between February 5, 2020 and January 27, 2021. February 5 is the day on which the first RT-PCR positive patient was diagnosed, a traveler arriving in Qatar [16].

The database included the swab date, SARS-CoV-2 laboratory result, and demographic information, including age, sex, and nationality. Age was categorized into 10-year age brackets, except for the last category (<10, 10-19, …, 80+). Nationality comprised 11 classifications. Nationalities with <1% of diagnosed SARS-CoV-2 cases were grouped as “Other nationalities”; nationalities with >1% of diagnosed cases constituted individual categories.

Two risk scores were derived. The “original” Qatar COVID-19 risk score was derived in April 2020, during the expanding phase of the epidemic [11], utilizing half of the RT-PCR tests administered from February 5, 2020 to April 21, 2020. This half of the sample was chosen randomly. Performance of the risk score was subsequently assessed and validated utilizing the remaining half of the sample.

Similarly, an updated version of the Qatar COVID-19 risk score was derived utilizing half of the RT-PCR testing sample compiled from February 5, 2020 to January 27, 2021. Performance of the updated risk score was subsequently assessed and validated utilizing the remaining half of the sample.

### Laboratory methods

For each individual, nasopharyngeal and/or oropharyngeal swabs (Huachenyang Technology, China) were collected for PCR testing and placed in Universal Transport Medium (UTM). Aliquots of UTM were extracted on the QIAsymphony platform (QIAGEN, USA) and tested with real-time reverse-transcription PCR (RT-qPCR) using TaqPath™COVID-19 Combo Kits (Thermo Fisher Scientific, USA) on ABI 7500 FAST (Thermo Fisher, USA). Samples were extracted using a custom protocol [25] on a Hamilton Microlab STAR (Hamilton, USA) and tested using AccuPower SARS-CoV-2 Real-Time RT-PCR Kit (Bioneer, Korea) on ABI 7500 FAST, or loaded directly into a Roche cobas® 6800 system and assayed with a cobas® SARS-CoV-2 Test (Roche, Switzerland). The first assay targets the viral S, N, and ORF1ab regions. The second targets the virus’ RdRp and E-gene regions, and the third targets the ORF1ab and E-gene regions. All testing was conducted following standardized protocols.

### Statistical analysis

#### Risk score derivation

Univariable logistic regressions were performed to identify associations between each demographic factor and SARS-CoV-2 status. Multivariable logistic regression was then conducted to identify independent predictors of SARS-CoV-2 RT-PCR positivity and to estimate adjusted odds ratios (aOR) and corresponding 95% confidence intervals (CI). A p-value ≤0.05 in the multivariable analysis for any predictor was considered to provide strong evidence for an association with the outcome. Predictors with p-values ≤0.05 were retained in deriving the Qatar COVID-19 risk score.

Each predictor level was assigned scoring points using the corresponding regression model’s β-coefficient multiplied by 10 (and rounded to the nearest integer) for ease of implementation, per established methodology [3-6, 26]. An aggregate risk score for each test was then derived by summing the scoring points, given the individual’s profile. No interaction terms between covariates were included, so as to keep the score accessible for broad use. The score was used to determine an individual’s level of risk of exposure to SARS-CoV-2 infection.

#### Risk score performance and validation

A receiving operating characteristics (ROC) curve was plotted to determine the capacity of the risk score to diagnose SARS-CoV-2 infection at different cut-off values resulting in a positive outcome. Sensitivity was defined as the proportion of those with a positive outcome when applying the score among tests with a positive RT-PCR result, that is, the capacity of the score to detect a true SARS-CoV-2 infection. Specificity was defined as the proportion of those with a negative outcome when applying the score among the tests with a negative RT-PCR result, that is, the capacity of the score to detect true absence of SARS-CoV-2 infection.

The optimal score cut-off/criterion to identify infected or uninfected cases was determined by selecting the value that maximized the sum of sensitivity and specificity. The area under the ROC curve (AUC) was also estimated to quantify the accuracy of the risk score, that is, how well the risk score separated infected from uninfected persons.

The risk score derived utilizing half of the sample was applied to the other half of the sample to assess and validate its performance. The risk score’s predictive and diagnostic performance was assessed by estimating the sensitivity, specificity, positive predictive value (PPV; probability of being infected given a positive outcome when applying the score), and the negative predictive value (NPV; probability of being uninfected given a negative outcome when applying the score).

#### Performance assessment of the original risk sore on prospective data

The “original” Qatar COVID-19 risk score that was derived from testing data up to April 21, 2020 was applied to all testing data from April 22, 2020 up to January 27, 2021. The diagnostic metrics described above were calculated to assess the performance of the risk score on data collected *after* its derivation.

Research methods were approved by the ethics review boards at Hamad Medical Center (HMC) and Weill Cornell Medicine-Qatar.

## Results

### Characteristics of SARS-CoV-2 RT-PCR testing conducted in Qatar

A total of 90,027 RT-PCR tests were performed between February 5, 2020 and April 21, 2020 for SARS-CoV-2 infection in Qatar, and 10,362 were positive, for an overall RT-PCR positivity of 11.5% (95% CI: 11.3%-11.7%). A total of 2,688,232 RT-PCR tests were performed between February 5, 2020 and January 27, 2021, and 200,646 proved positive for an overall RT-PCR positivity of 7.5% (95% CI: 7.4%-7.5%). Characteristics of SARS-CoV-2 RT-PCR testing conducted in Qatar are presented in Table 1.

**Table 1.** Characteristics of SARS-CoV-2 RT-PCR testing conducted in Qatar.

| Characteristics | Original sample of Feb 5, 2020-Apr 21, 2020 |  |  | Extended sample of Apr 22, 2020-Jan 27, 2021 |  |  |
| --- | --- | --- | --- | --- | --- | --- |
|  | Tested<br>N=90,027 | SARS-CoV-2 RT-PCR positive<br>N=10,362 (11.5%) |  | Tested<br>N=2,598,205 | SARS-CoV-2 RT-PCR positive<br>N=190,284 (7.3%) |  |
|  | N (%) | N (%) | P-value | N (%) | N (%) | P-value |
| Sex |  |  |  |  |  |  |
| Male | 69,440 (77.1) | 9,022 (13.0) | <0.001 | 1,773,809 (68.3) | 144,875 (8.2) | <0.001 |
| Female | 20,586 (22.9) | 1,339 (6.5) |  | 824,220 (31.7) | 45,393 (5.5) |  |
| Age (years) |  |  |  |  |  |  |
| <10 | 2,266 (2.5) | 175 (7.7) | <0.001 | 206,506 (7.9) | 12,806 (6.2) | <0.001 |
| 10-19 | 3,493 (3.9) | 221 (6.3) |  | 184,501 (7.1) | 11,606 (6.3) |  |
| 20-29 | 25,844 (28.7) | 2,594 (10.0) |  | 601,053 (23.1) | 43,073 (7.2) |  |
| 30-39 | 30,823 (34.2) | 3,578 (11.6) |  | 834,766 (32.1) | 64,301 (7.7) |  |
| 40-49 | 16,087 (17.9) | 2,173 (13.5) |  | 450,517 (17.3) | 36,374 (8.1) |  |
| 50-59 | 7,658 (8.5) | 1,085 (14.2) |  | 219,455 (8.4) | 15,715 (7.2) |  |
| 60-69 | 2,791 (3.1) | 403 (14.4) |  | 77,662 (3.0) | 4,973 (6.4) |  |
| 70-79 | 737 (0.8) | 115 (15.6) |  | 18,109 (0.7) | 1,090 (6.0) |  |
| 80+ | 328 (0.4) | 18 (5.5) |  | 5,635 (0.2) | 346 (6.1) |  |
| Nationality |  |  |  |  |  |  |
| Other* | 13,448 | 909 (6.8) | <0.001 | 646,379 (24.9) | 26,124 (4.0) | <0.001 |
| Bangladeshi | 8,622 | 2,013 (23.3) |  | 142,649 (5.5) | 18,932 (13.3) |  |
| Nepalese | 8,024 | 1,712 (21.3) |  | 187,623 (7.2) | 26,393 (14.1) |  |
| Indian | 17,232 | 2,632 (15.3) |  | 557,106 (21.4) | 49,085 (8.8) |  |
| Pakistani | 4,036 | 643 (15.9) |  | 135,337 (5.2) | 10,736 (7.9) |  |
| Kenyan | 909 | 110 (12.1) |  | 36,505 (1.4) | 2,114 (5.8) |  |
| Egyptian | 2,836 | 298 (10.5) |  | 140,872 (5.4) | 9,808 (7.0) |  |
| Sri Lankan | 2,281 | 174 (7.6) |  | 55,635 (2.1) | 6,450 (11.6) |  |
| Sudanese | 2,058 | 166 (8.1) |  | 83,714 (3.2) | 5,222 (6.2) |  |
| Filipino | 6,408 | 352 (5.5) |  | 185,033 (7.1) | 13,171 (7.1) |  |
| Qatari | 24,173 | 1,353 (5.6) |  | 427,352 (16.4) | 22,249 (5.2) |  |

RT-PCR; real-time polymerase chain reaction
\*These include 148 other nationalities residing in Qatar.

### Original Qatar COVID-19 risk score

#### Risk score derivation

Univariable logistic regression of half the testing sample from February 5, 2020-April 21, 2020 identified significant associations between individual variables, age, sex, and nationality, and RT-PCR outcome (Table 2A). All three demographic variables were retained in the multivariable logistic regression and were included in the risk score. Scoring points were lower for females than for males and higher for specific nationalities. The risk score was expressed as a mathematical formula illustrated in Box 1A.

**Table 2.** Results of regression analyses used to derive a) the original and b) updated Qatar COVID-19 risk scores.

**A) “Original” Qatar COVID-19 risk score**
| Characteristics | “Original” Qatar COVID-19 risk score |  |  |
| --- | --- | --- | --- |
| | $\beta$ | aOR (95% CI) | Score points |
| Sex |  |  |  |
| Male | 0.000 | 1.00 | 0 |
| Female | -0.384 | 0.68 (0.62-0.75) | -4 |
| Age (years) |  |  |  |
| <10 | 0.000 | 1.00 | 0 |
| 10-19 | -0.028 | 0.97 (0.72-1.31) | 0 |
| 20-29 | -0.123 | 0.88 (0.70-1.12) | -1 |
| 30-39 | -0.109 | 0.90 (0.71-1.13) | -1 |
| 40-49 | 0.100 | 1.11 (0.87-1.40) | 1 |
| 50-59 | 0.295 | 1.34 (1.05-1.72) | 3 |
| 60-69 | 0.561 | 1.75 (1.34-2.30) | 6 |
| 70-79 | 1.030 | 2.80 (1.97 (3.99) | 10 |
| 80+ | -0.083 | 0.92 (0.45-1.88) | -1 |
| Nationality |  |  |  |
| Other* | 0.000 | 1.00 | 0 |
| Bangladeshi | 1.362 | 3.91 (3.45-4.93) | 14 |
| Nepalese | 1.358 | 3.89 (3.43-4.41) | 14 |
| Indian | 0.897 | 2.45 (2.19-2.75) | 9 |
| Pakistani | 0.852 | 2.35 (2.01-2.74) | 9 |
| Kenyan | 0.902 | 2.47 (1.85-3.28) | 9 |
| Egyptian | 0.459 | 1.58 (1.30-1.93) | 5 |
| Sri Lankan | 0.059 | 1.06 (0.83-1.35) | 1 |
| Sudanese | 0.318 | 1.37 (1.09-1.74) | 3 |
| Filipino | -0.083 | 0.92 (0.77-1.11) | -1 |
| Qatari | -0.205 | 0.82 (0.72-0.92) | -2 |

B) “Updated” Qatar COVID-19 risk score
| Characteristics | “Updated” Qatar COVID-19 risk score |  |  |
| --- | --- | --- | --- |
| | $\beta$ | aOR (95% CI) | Score points |
| Sex |  |  |  |
| Male | 0.000 | 1.00 | 0 |
| Female | -0.193 | 0.82 (0.81-0.84) | -2 |
| Age (years) |  |  |  |
| <10 | 0.000 | 1.00 | 0 |
| 10-19 | 0.077 | 1.08 (1.04-1.12) | 1 |
| 20-29 | -0.160 | 0.85 (0.83-0.88) | -2 |
| 30-39 | -0.088 | 0.92 (0.89-0.94) | -1 |
| 40-49 | -0.017 | 1.02 (0.99-1.05) | 0 |
| 50-59 | -0.017 | 0.98 (0.95-1.02) | 0 |
| 60-69 | -0.028 | 0.97 (0.93-1.02) | 0 |
| 70-79 | 0.049 | 1.05 (0.96-1.15) | 0 |
| 80+ | 0.068 | 1.07 (0.91-1.26) | 1 |
| Nationality |  |  |  |
| Other* | 0.000 | 1.00 | 0 |
| Bangladeshi | 1.307 | 3.69 (3.59-3.80) | 13 |
| Nepalese | 1.368 | 3.93 (3.83-4.03) | 14 |
| Indian | 0.830 | 2.29 (2.26-2.35) | 8 |
| Pakistani | 0.718 | 2.05 (1.99-2.12) | 7 |
| Kenyan | 0.478 | 1.61 (1.51-1.72) | 5 |
| Egyptian | 0.553 | 1.74 (1.68-1.80) | 6 |
| Sri Lankan | 1.096 | 2.99 (2.87-3.12) | 11 |
| Sudanese | 0.456 | 1.58 (1.51-1.65) | 5 |
| Filipino | 0.647 | 1.91 (1.85-1.97) | 6 |
| Qatari | 0.236 | 1.27 (1.23-1.30) | 2 |

$\beta$ , beta coefficient; aOR, adjusted odds ratio; CI, confidence interval.
\*These include 148 other nationalities residing in Qatar.

##### Box 1. Mathematical formula for the A) original and the B) updated Qatar COVID-19 risk scores.

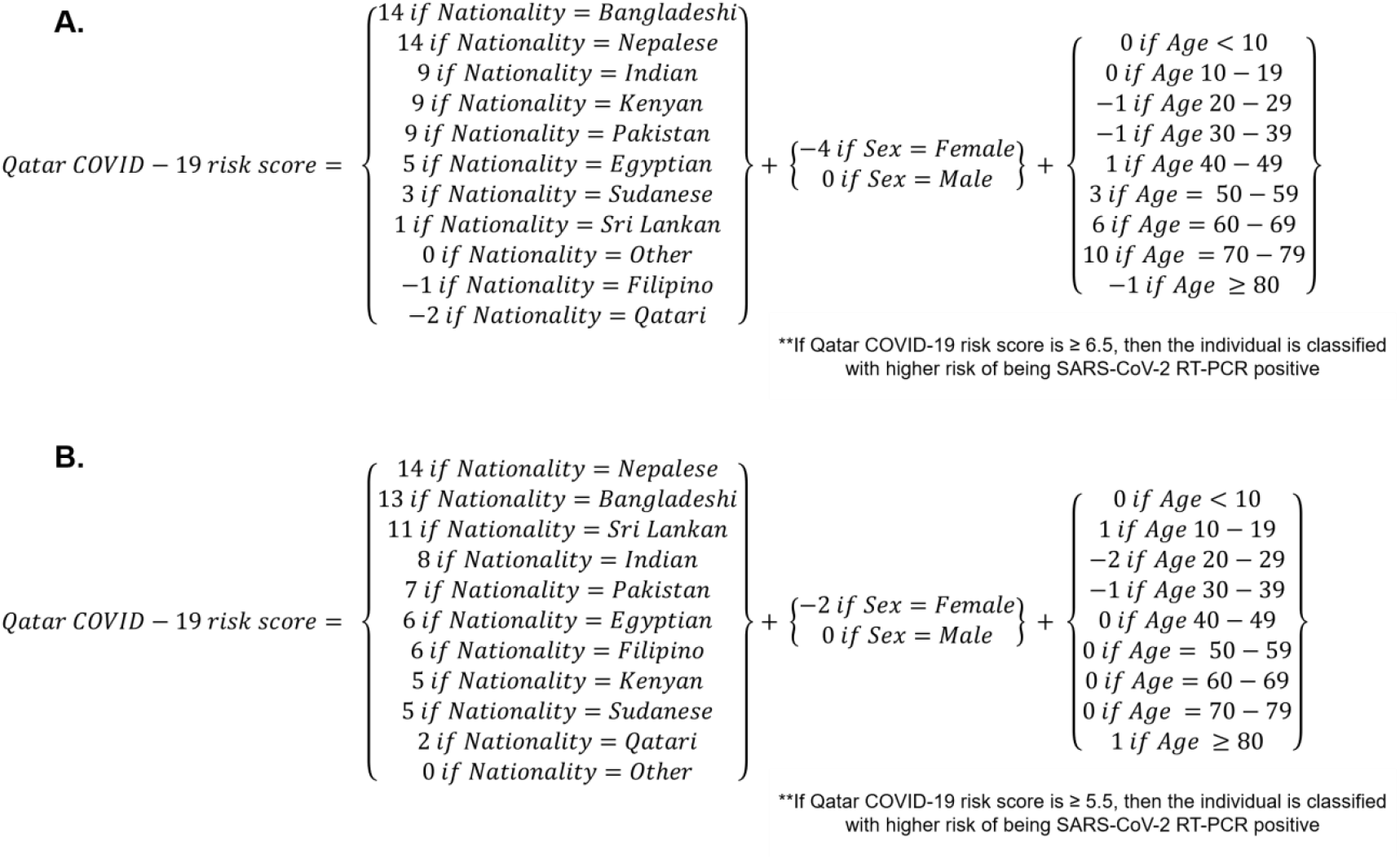

#### Risk score performance and validation

The ROC curve was generated, and the AUC was estimated at 0.67 (95% CI: 0.66-0.67) (Figure 1A). A score cut-off value of 6.5 maximized the sum of sensitivity and specificity. This indicated that individuals with a risk score ≥6.5 should be prioritized for RT-PCR testing.

**Figure 1.**
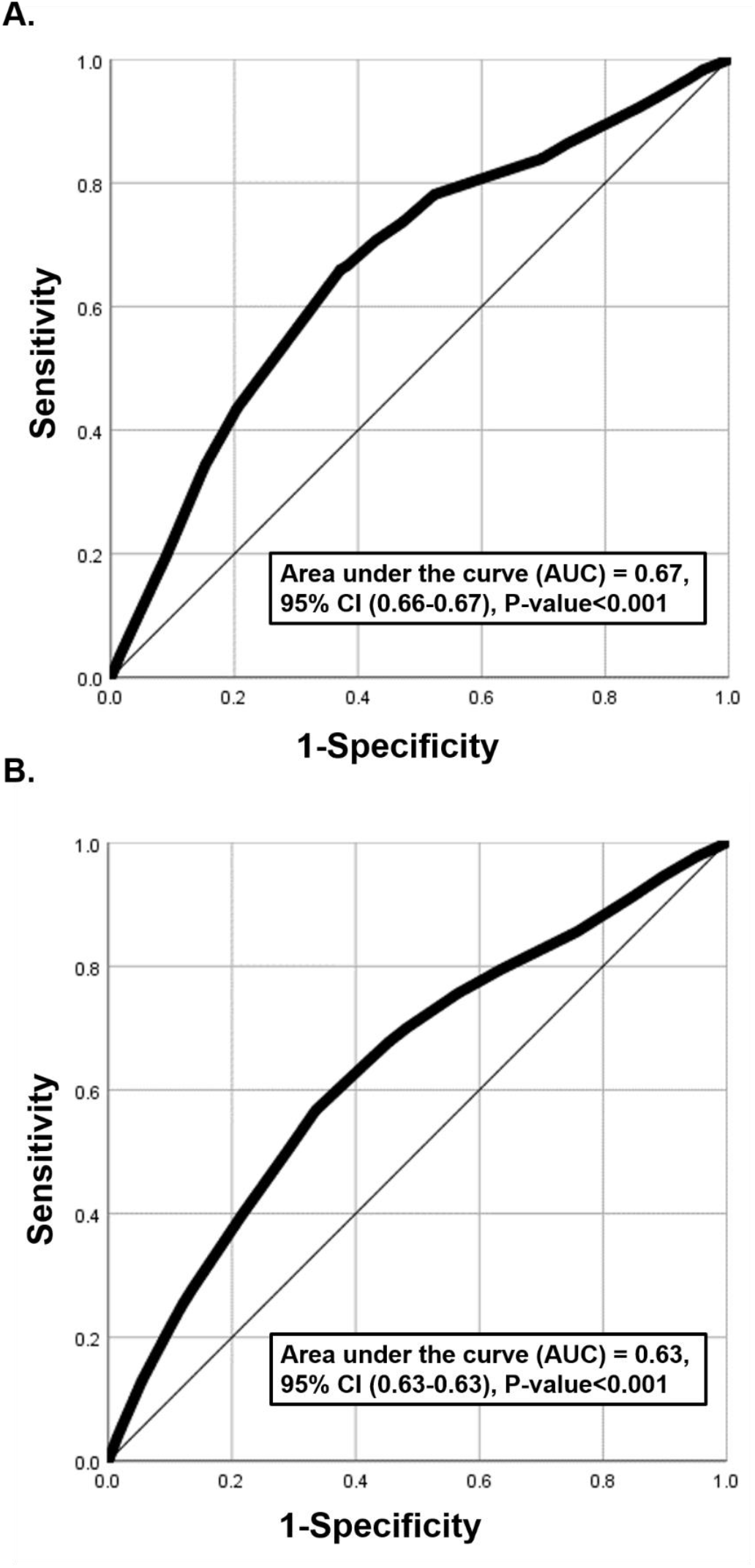
Diagnostic performance of the A) original and B) updated Qatar COVID-19 risk scores, assessed using the area under the receiver operating characteristic (ROC) curve.

To validate the risk score, it was applied to the other half of the sample and yielded the following diagnostic metrics: 66.8% (95% CI: 65.5%-68.0%) for sensitivity, 62.6% (95% CI: 62.2%-63.1%) for specificity, 19.1% (95% CI: 18.6%-19.7%) for PPV, and 93.4% (93.1%-93.7%) for NPV (Table 3).

**Table 3.** Validation and diagnostic performance of the Qatar COVID-19 risk score, assessed using measures of sensitivity, specificity, positive predictive value, and negative predictive value.

|  |  | SARS-CoV-2 infection status using RT-PCR |  |  | Risk score metrics |  |  |  |
| --- | --- | --- | --- | --- | --- | --- | --- | --- |
|  |  | Positive | Negative | Total | Sensitivity | Specificity | Positive Predictive Value | Negative Predictive Value |
|  |  |  |  |  | % (95% CI) | % (95% CI) | % (95% CI) | % (95% CI) |
| **SARS-CoV-2 infection status using the “original” Qatar COVID-19 risk score applied to the original sample of Feb 5, 2020-Apr 21, 2020** | Positive | 3,520 | 14,876 | 18,396 | 66.8<br>(65.5-68.0) | 62.6<br>(62.2-63.1) | 19.1<br>(18.6-19.7) | 93.4<br>(93.1-93.7) |
|  | Negative | 1,752 | 24,951 | 26,703 |  |  |  |  |
|  | Total | 5,272 | 39,827 | 45,099 |  |  |  |  |
| **SARS-CoV-2 infection status using the “updated” Qatar COVID-19 risk score applied to the updated sample of Feb 5, 2020-Jan 27, 2021** | Positive | 58,735 | 482,098 | 540,833 | 59.4<br>(59.1-59.7) | 61.1<br>(61.1-61.2) | 10.9<br>(10.8-10.9) | 94.9<br>(94.9-95.0) |
|  | Negative | 40,168 | 758,474 | 798,642 |  |  |  |  |
|  | Total | 98,903 | 1,240,572 | 1,339,475 |  |  |  |  |
| **SARS-CoV-2 infection status using the “original” Qatar COVID-19 risk score by applying it prospectively to the extended sample of Apr 22, 2020-Jan 27, 2021** | Positive | 100,183 | 836,863 | 937,046 | 52.6<br>(52.4-52.9) | 65.2<br>(65.2-65.3) | 10.7<br>(10.6-10.8) | 94.6<br>(94.6-94.6) |
|  | Negative | 90,101 | 1,571,058 | 1,661,159 |  |  |  |  |
|  | Total | 190,284 | 2,407,921 | 2,598,205 |  |  |  |  |

\*RT-PCR, real-time polymerase chain reaction.

### Updated Qatar COVID-19 risk score

#### Risk score derivation

Univariable logistic regression of half the testing sample from April 22, 2020-January 27, 2021 identified significant associations between individual variables, age, sex, and nationality and RT-PCR outcome (Table 2B). All three demographic variables were retained in the multivariable logistic regression and were included in the risk score. Scoring points were lower for females than for males and higher for specific nationalities. The risk score was expressed as a mathematical formula illustrated in Box 1B.

#### Risk score performance and validation

The ROC curve was generated, and the AUC was estimated at 0.63 (95% CI: 0.63-0.63) (Figure 1B). A score cut-off value of 5.5 maximized the sum of sensitivity and specificity. This indicated that individuals with risk scores ≥5.5 should be prioritized for RT-PCR testing.

To validate the risk score, it was applied to the other half of the sample and yielded the following diagnostic metrics: 59.4% (95% CI: 59.1%-59.7%) for sensitivity, 61.1% (95% CI: 61.1%-61.2%) for specificity, 10.9% (95% CI: 10.8%-10.9%) for PPV, and 94.9% (94.9%-95.0%) for NPV (Table 3).

### Performance assessment of the original derived risk sore on prospective data

The original Qatar COVID-19 risk score derived from testing data until April 21, 2020 was applied to all testing data from April 22, 2020 to January 27, 2021. It yielded the following diagnostic metrics: 52.6% (95% CI: 52.4%-52.9%) for sensitivity, 65.2% (95% CI: 65.2%-65.3%) for specificity, 10.7% (95% CI: 10.6%-10.8%) for PPV, and 94.6% (94.6%-94.6%) for NPV (Table 3).

## Discussion

To illustrate the concept and public health value of COVID-19 risk scores, we derived a COVID-19 risk score for Qatar, which to our knowledge is the first such score for any country. Its relatively strong performance and high diagnostic metrics supported the utility of using them to inform national testing strategies. The score provided a non-invasive tool for identification of individuals at higher risk of being infected, who should be prioritized for RT-PCR testing, in addition to typical cases of clinical suspicion and contact tracing. Therefore, use of such scores may substantially enhance the effectiveness of the “testing, tracing, and isolation” approach that is currently the “backbone” of COVID-19 national response in different countries [2]. Indeed, the present analyses have helped to guide Qatar’s national COVID-19 response to control transmission and to reduce the disease burden.

A main finding of the present study is that the COVID-19 risk score performed similarly to other public health risk scores, such as those for diabetes [6, 26-31]. Indeed, this risk score, though simple to implement, demonstrated reasonably high diagnostic accuracy (Figure 1 and Table 3). The original risk, which was derived based on early epidemic data until only April, 2020 proved effective and offered comparable performance to the updated risk score based on all data until the present (Table 3). This further affirms the utility of such scores even when they are derived from a more limited set of testing data during a specific phase of the epidemic.

While our study provided a proof of concept for the use of such scores, implementation of them can be further optimized. We reported a risk score derived over one year. The score’s performance could have been improved, with higher diagnostic ability, if different scores were derived in real-time at every phase of the epidemic and their use is updated continuously. It is remarkable that the risk score derived using a year of RT-PCR testing performed well, even though the epidemiology of the infection in Qatar has evolved immensely during this year [11, 14-24]. A month-by-month risk score, derived based on RT-PCR testing of only the previous month, would have better predicted the risk of infection month by month. With the ease of the process of deriving such risk scores, continuous updating of risk scores is feasible even in resource-limited settings, provided there is a minimal digital healthcare system to track RT-PCR testing.

A finding of this study is that there is always likely to be considerable variation in the risk of exposure to the infection based on basic demographics (such as age, sex, and nationality). This reflects the underlying dynamics of infection transmission in any country, as those delineated earlier for Qatar [11, 14-24]. Biological factors such as age [32-38], may also cause variation in susceptibility to the infection or in the likelihood of the infection’s being symptomatic, which may affect the likelihood of testing or of a positive test outcome. A COVID-19 risk score can be seen as a metric that quantifies these variations in any setting, creating an opportunity for more effective public health action that addresses the needs of different segments of the population.

This study has some limitations. The COVID-19 risk score was derived using the national testing database rather than a nationally representative, probability-based survey of the total population of Qatar. Infection levels and patterns among tested individuals may not necessarily reflect actual levels and patterns in the wider population. The score used a small number of demographic variables, but its predictive power might have been enhanced if other variables had been available, such as more socio-demographic indicators. Despite these limitations, the study had important strengths. The testing database encompassed all RT-PCR testing done in Qatar up the present and was massive, including results of over two million tests, representing a majority of the population of Qatar [8, 39]. While other variables in the score might have improved its predictive power, they might have reduced its accessibility and utility for broad use as a tool of public health.

In conclusion, the concept and utility of a COVID-19 risk score was demonstrated in a single country. Such public health tool, based on a set of non-invasive and easily captured variables, can help to optimize testing and suppression of infection transmission, while maximizing efficient use of available resources.

## Data Availability

All relevant data are available within the manuscript.

## Acknowledgments

We thank Her Excellency Dr. Hanan Al Kuwari, Minister of Public Health, for her vision, guidance, leadership, and support. We also thank Dr. Saad Al Kaabi, Chair of the System Wide Incident Command and Control (SWICC) Committee for the COVID-19 national healthcare response, for his leadership, analytical insights, and for his instrumental role in implementing data information systems that made these studies possible. We further extend our appreciation to SWICC Committee and Scientific Reference and Research Taskforce (SRRT) members for their informative input, scientific technical advice, and enriching discussions. We also thank Dr. Mariam Abdulmalik, CEO of the Primary Health Care Corporation and the Chairperson of the Tactical Community Command Group on COVID-19, as well as members of this committee, for providing support to the teams that worked on the field surveillance. We further thank Dr. Nahla Afifi, Director of Qatar Biobank (QBB), Ms. Tasneem Al-Hamad, Ms. Eiman Al-Khayat and the rest of the QBB team for their unwavering support in retrieving and analyzing samples and in compiling and generating databases for COVID-19 infection, as well as Dr. Asmaa Al-Thani, Chairperson of the Qatar Genome Programme Committee and Board Vice Chairperson of QBB, for her leadership of this effort. We also acknowledge the dedicated efforts of the Clinical Coding Team and the COVID-19 Mortality Review Team, both at Hamad Medical Corporation, and the Surveillance Team at the Ministry of Public Health.

## Author contributions

LJA conceived and designed this study. SRD and HC contributed to study design and conducted the statistical analyses. SRD and LJA wrote the first draft of the article. All authors contributed to development of the study protocol, data collection, database development, discussions and interpretation of the results, and to the writing of the manuscript. All authors have read and approved the final manuscript.

